# A Heterogeneous Graph Neural Network Framework for Multi-Horizon Stroke Mortality Prediction

**DOI:** 10.64898/2026.06.09.26355176

**Authors:** Aabila Tharzeen, Alireza Vafaei Sadr, Nazli Radfar, Wenke Hwang, Vida Abedi, Ramin Zand

## Abstract

**Background:** Machine learning models for stroke mortality prediction typically treat each time horizon independently and use flat tabular features that ignore the relational structure of electronic health records (EHRs). In this pilot study, we leveraged graph-based machine learning models to predict post stroke all-cause-mortality across three different time horizons.

**Methods:** We developed Stroke Temporal Heterogeneous Graph (StrokeTHG), a heterogeneous graph neural network model for simultaneous multi-horizon stroke mortality prediction (30-day, 90-day, 1-year) using EHR data from Penn State Health System. The model encodes various relations among EHR entities (e.g., patient, diagnosis, comorbidity) and temporal encoding of admission time to better predict stroke mortality. We compared our proposed approach against various baseline methods, including Logistic Regression, Random Forest, and XGBoost. We also performed ablation and subgroup analyses, evaluated the quality of learned graph embeddings, and assessed the importance of different edge types in the graph.

**Results:** We included 4,144 stroke patients (mean age 69.2 years; 54.3% men), of whom 3,332 (80.4%) survived their stroke after one year. 30-day, 90-day, and 1-year mortality rates were 9.7%, 13.7%, and 19.6%, respectively. Our proposed approach, StrokeTHG, achieved AUROC of 0.872, 0.878, and 0.837 across horizons, outperforming all tabular baselines. At ≥75% specificity, the model identified 5–10 percentage points more mortality cases than the best baseline at each horizon. Subgroup analysis demonstrated consistent performance across sex subgroups and the largest discriminative gains in the Age 65–80 stratum. Edge-type ablation identified phenotype–patient and admission–patient edges in the constructed EHR graph as the most influential relational edges for mortality prediction. StrokeTHG embeddings outperformed all graph and matrix factorization baselines under an identical downstream classifier, confirming that performance gains stem from representation quality rather than classifier capacity.

**Conclusions:** StrokeTHG demonstrates that heterogeneous graph representations of EHR data provide a consistent improvement over flat tabular models for multi-horizon stroke mortality prediction, with particular advantage at clinically actionable sensitivity thresholds and novel multi-horizon monotonic prediction capability. This methodological framework may be adaptable to other EHR-based clinical research studies seeking to leverage heterogeneous relational structures for predictive modeling.

## Introduction

Post-stroke mortality prediction at multiple time horizons can help clinicians personalize care, guide prognosis-based decision-making, and allocate healthcare resources more effectively^1–3^. Prior machine learning approaches to stroke mortality prediction have various limitations. First, existing approaches often treat each prediction window independently^4–6^. Second, they typically represent patient records as flat feature vectors, thereby discarding the rich relational structure encoded in electronic health record (EHR) data^7,8^. Connections among patients, diagnoses, comorbidities, facilities, providers, and clinical phenotypes may contain clinically meaningful signals that tabular models cannot fully exploit^5,9^.

Graph neural networks (GNNs) offer a principled framework for modeling the relational structure of EHRs, including links among patients, encounters, diagnoses, comorbidities, providers, facilities, and clinical phenotypes.^10–12^. The Temporal Heterogeneous Graph neural networks ^13,14^extend this approach by learning relation-specific representations across multiple node and edge types, making it naturally suited to the heterogeneous entity structure of clinical data.

In this pilot study, we leverage heterogeneous graph^15^ principles and propose StrokeTHG, a heterogeneous graph framework for simultaneous 30-day, 90-day, and 1-year post-stroke mortality prediction. StrokeTHG models inherent EHR relationships and incorporates monotonic prediction heads that jointly estimate mortality risk across all three horizons while enforcing coherent probability ordering: P_mortality_ (30 day) ≤ P_mortality_ (90 day) ≤ P_mortality_ (1 year). We further benchmark StrokeTHG against traditional tabular machine learning models, including logistic regression, random forest, and XGBoost^16–18^, to assess whether heterogeneous EHR graph representations improve multi-horizon mortality prediction beyond conventional feature-vector methods. Beyond stroke mortality prediction, this framework may provide a generalizable approach for other EHR-based research studies in which heterogeneous relationships among patients, clinical entities, and healthcare processes contain meaningful predictive signals.

## Methods

### Study design and Data source

We conducted a retrospective cohort study using electronic health record (EHR) data from Penn State Health (PSH), a multi-hospital academic health system in Pennsylvania, from 2017 to 2023. PSH contributed approximately 20.3 million raw EHR records for the stroke cohort, including 7,433 patient demographic records, 3,968,637 diagnosis records, 442,729 encounter records, and 10,035,665 laboratory result records.

The PSH cohort was constructed using a registry-based phenotyping pipeline. Patients were identified through the institutional stroke registry, matched to encounter records, and further filtered according to the following criteria: primary discharge diagnosis of ischemic stroke (ICD-9 and ICD-10 codes provided in Supplemental Material); inpatient or emergency encounter type; and encounter duration of at least 24 hours. Mortality outcomes were ascertained from EHR death records and National Death Index (NDI) linkage.

The final analysis cohort included 4,144 patients, each represented by a single index stroke encounter. Patients were 18 years of age or older at the index stroke encounter. The cohort was split into training, validation, and test sets using a 60/20/20 ratio, stratified by 30-day mortality to ensure representative class distributions across partitions. Missing values were imputed using Pympute ^19^, an EHR-focused imputation toolkit designed to handle missingness in clinical data. Pairwise statistical significance of AUROC differences between StrokeTHG and each baseline was assessed using paired bootstrap hypothesis testing (10000 resamples, two-sided) with 95% confidence intervals for AUROC differences.

### Outcome Definition

Three binary all-cause mortality outcomes were defined from the index stroke discharge date: (1) 30-day mortality; (2) 90-day mortality; and (3) 1-year mortality. These horizons were selected to capture the acute (30-day), subacute rehabilitation (90-day), and chronic secondary prevention (1-year) clinical decision timeframes. Mortality follow-up was measured from the index stroke discharge date to align prediction with post-discharge risk stratification.

### Heterogeneous Graph Construction

We constructed a heterogeneous graph with nine node types and 19 edge types encoding all clinically relevant relationships in the EHR. Node and edge types were selected to reflect clinically meaningful relationships that are routinely available in structured health system data, including patient demographics, encounters, diagnoses, comorbidities, facilities, providers, care pathways, disease-severity phenotypes, and patient-similarity patterns. This design was intended to preserve both individual-level clinical information and higher-order relational context.

The nine node types were: (1) Patient nodes; (2) Admission nodes encoding encounter-level features (3) Diagnosis nodes representing ICD-10 code groups; (4) Facility nodes encoding hospital identifiers; (5) Provider nodes encoding specialty type; (6) Comorbidity nodes representing chronic condition categories; (7) Phenotype nodes representing lab-derived severity cluster centroids computed by K-means clustering on 42 laboratory values, serving as implicit severity anchors; (8) Clinical pathway nodes represented admitting pathway assignments. (9) DRG tier nodes represented Medicare Severity DRG complexity categories.

The 19 edge types included (Table 1): standard patient-encounter-diagnosis-facility-provider-comorbidity-phenotype-pathway-DRG relational edges; ICD-10 ontology hierarchy edges (12,884 edges encoding L1 block, L2 chapter, and L3 stroke subtype relationships); and two patient-to-patient K-nearest-neighbor similarity^20^ channels: a broad KNN (K=25, cosine similarity over all patient features, 165,432 edges) and a laboratory-specific KNN (K=15, cosine similarity over 42 laboratory values only, 100,868 edges). The dual KNN design allows the model to aggregate signals from patients who are similar overall and from those who are specifically similar based on laboratory-value profiles. Both KNN graphs were constructed by connecting each patient exclusively to their K nearest training-set neighbors, ensuring no direct similarity edges exist between validation or test patients. Mortality outcomes and follow-up information were not used to construct node features or any other graph edges.

**Table 1:**
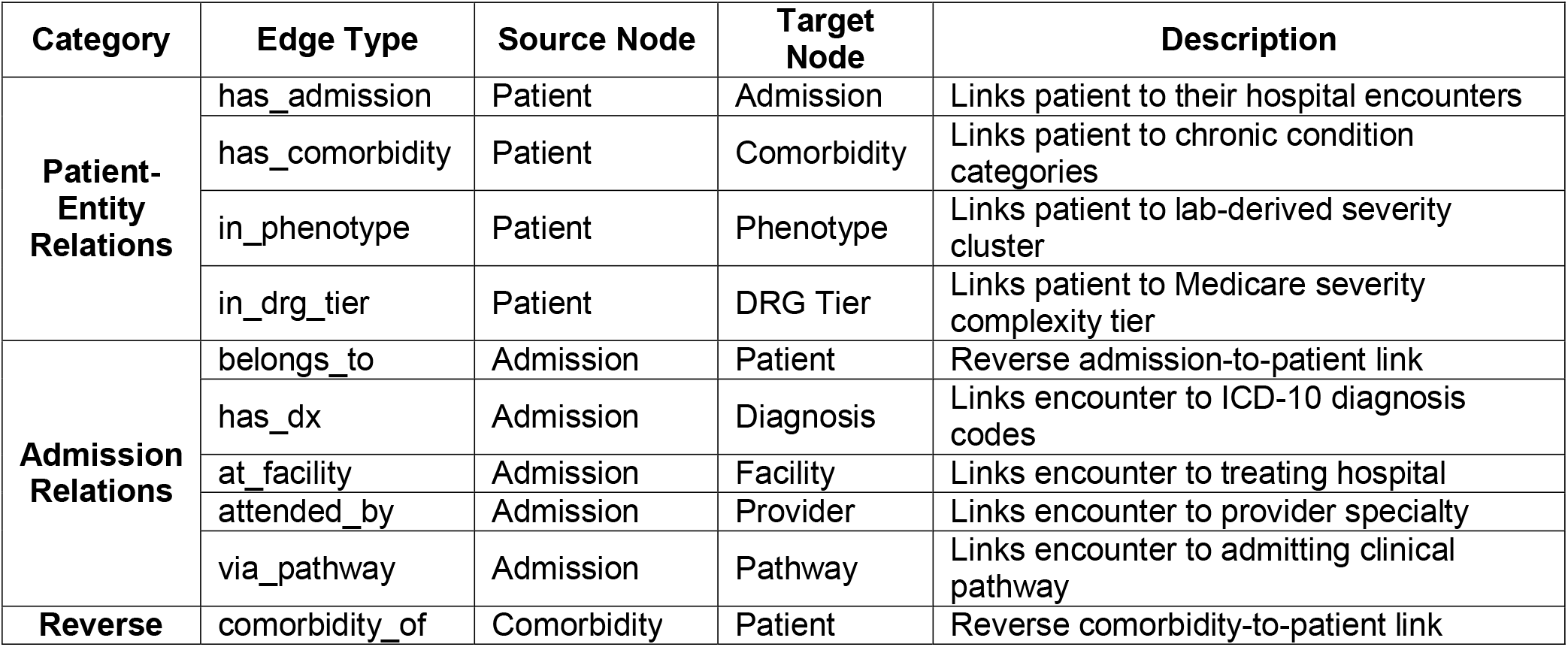
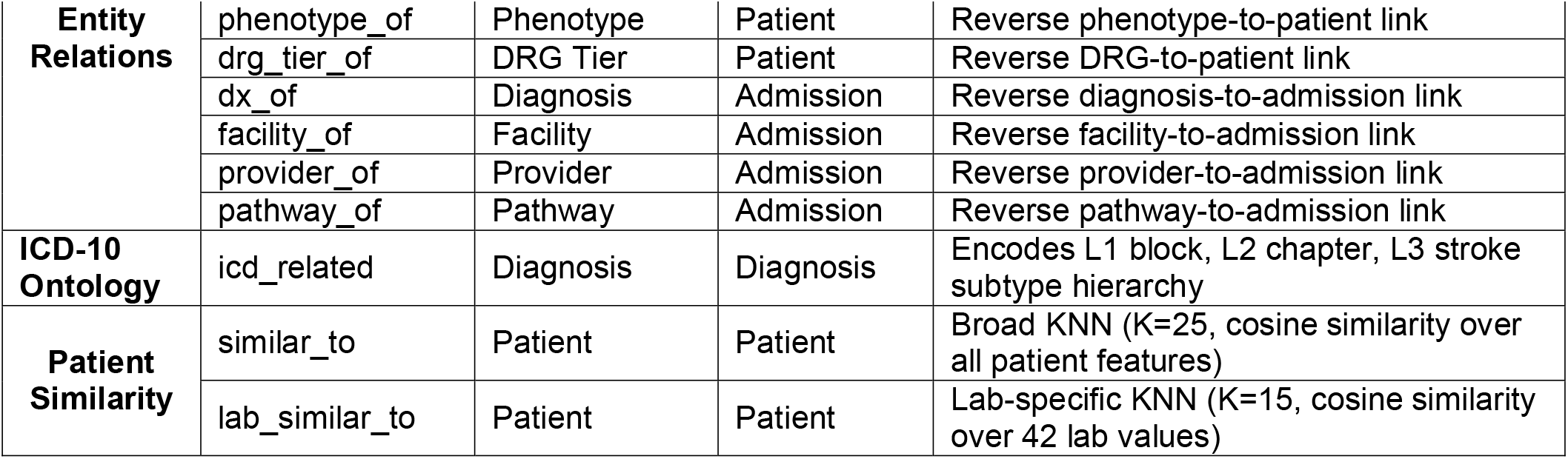
Edge Types in the StrokeTHG.

### StrokeTHG Architecture

StrokeTHG includes five main components. First, node features from each node type are projected into a 128-dimensional hidden space using node-type-specific linear layers followed by layer normalization and GELU activation^21^. Second, the model uses three relational message-passing layers, each applying edge-type-specific Multilayer Perceptron (MLP) transforms with learned relation gates and scatter-mean aggregation, followed by layer normalization, 0.35 dropout, and stochastic depth of 0.05 with residual connections. Third, admission time information is incorporated using sinusoidal temporal embeddings, which are gated and added to admission node representations prior to message passing. Fourth, patient-level graph embeddings are concatenated with tabular patient features and passed through a learned projection layer, allowing the model to balance information from the graph structure and structured clinical variables. Finally, three monotonic prediction heads jointly estimate 30-day, 90-day, and 1-year mortality risk. The monotonic risk head produces three outputs via:

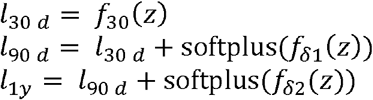

Where *f*_30_ (*z*), *f*_*δ*1_ (z) and *f*_*δ*2_ (*z*) are MLP blocks and z is the patient embedding and predicted mortality probability at each horizon is obtained via P(mortality) = sigmoid(*l*). Since softplus^22^ outputs are strictly positive, this formulation guarantees *P*_30_ ≤ *P*_90_ ≤ *P*_1_*y* by construction, ensuring that shorter-term risk does not exceed longer-term risk.

### Model Training and baseline methods

We first performed masked feature pre-training for 150 epochs, during which 30% of patient and admission node features were randomly masked, and the model was trained to reconstruct them from the surrounding graph structure. For supervised training, StrokeTHG was trained transductively, the full graph was loaded for message passing across all nodes, with loss computed exclusively over training-set patients using binary masks; validation and test patient labels were not used for gradient updates; validation labels were used only for model selection and early stopping, while test labels were used only for final performance evaluation. This transductive setup is consistent with standard practice in graph learning literature ^23^, and does not constitute label leakage: the cohort comprises a single index encounter per patient with no longitudinal follow-up visits, and outcome labels from validation and test patients are never used in any computation during training. KNN patient similarity edges were constructed in a leakage-safe manner, connecting each patient exclusively to their K nearest training-set neighbors, ensuring no direct edges exist between validation or test patients through similarity channels. We used the AdamW optimizer (learning rate 3 × 10^−4^, weight decay 10^−4^) with cosine annealing with warm restarts (T_0_=100). Training ran for up to 500 epochs with early stopping based on mean validation AUROC across all three horizons, evaluated every 5 epochs with a patience of 60 evaluations. To address class imbalance, we used asymmetric focal loss (γ_+_=1.0, γ_−_=3.0, label smoothing=0.05) with dynamic task weighting to balance learning across the three prediction horizons. Gradient norms were clipped at 2.0. All experiments used a single random seed (42) for reproducibility; confidence intervals were estimated using Bias-Corrected and Accelerated (BCa) bootstrap resampling (500 iterations) applied to held-out test set predictions. Missing values were imputed using Pympute^19^, an EHR-focused imputation toolkit; no missingness indicators were included. Continuous features were standardized using parameters estimated from the training set only to prevent data leakage. Baseline models: Logistic Regression, Random Forest, and XGBoost, were trained inductively using hyperparameters tuned on validation set on the same 253 tabular features and identical train/validation/test split as StrokeTHG. All models were implemented in Python using PyTorch 2.8.0 and PyTorch Geometric 2.5.2^24^, with scikit-learn^25^ for baseline models.

## Results

### Patient characteristics

A total of 4,144 patients met the inclusion criteria and formed the final analysis cohort. The mean age at index stroke was 69.0 years (SD 14.7, range 18–89) and 54.3% were men (n=2,252). The cohort was predominantly White (83%), with Black/African American (6.7%), Multiracial (4.7%), and Asian (1.7%). All included patients had ischemic stroke as their primary discharge diagnosis. Stroke mortality rates were 9.7% (n=403) at 30 days, 13.7% (n=568) at 90 days, and 19.6% (n=812) at one year. The three most common comorbidities were hypertension, dyslipidemia, and diabetes mellitus, with patients carrying an average of 4.1 comorbid conditions per encounter. The supplemental material presents the full demographic and clinical characteristics of the cohort.

### Discriminative Performance Analysis

We evaluated StrokeTHG against three tabular baseline classifiers on a held-out test set of PSH stroke patients across three mortality prediction horizons post-stroke. Model performance was assessed using AUROC, AUPRC, sensitivity, and specificity,^26^ with classification thresholds determined via the Youden index^27^. BCa bootstrap 95% confidence intervals were computed for all metrics. StrokeTHG achieved the highest AUROC and AUPRC at all three horizons compared to tabular baselines, with the most pronounced advantages in sensitivity and precision-recall trade-offs at shorter horizons. Paired bootstrap hypothesis testing (10,000 resamples) confirmed statistically significant AUROC improvements for StrokeTHG in 6 of 9 pairwise comparisons (p<0.05), with the strongest evidence at the 90-day horizon where all three baseline comparisons reached significance. No baseline outperformed StrokeTHG in any comparison. Full pairwise results are reported in the Supplemental Material.

The ROC curves for all the models across various horizons are shown in Figure 1. StrokeTHG ranked first at all three prediction horizons on the PSH test set. At 30 days (Figure 2), the model achieved AUROC 0.872 (95% CI 0.814–0.917) and AUPRC 0.393, outperforming the next best baseline XGBoost (AUROC 0.832, AUPRC 0.336). At 90 days (Figure 2), StrokeTHG achieved AUROC 0.878 (95% CI 0.826–0.912) and AUPRC 0.511, surpassing XGBoost (AUROC 0.823, AUPRC 0.498) and all other baselines. At one year (Figure 3), AUROC was 0.837 with AUPRC 0.542, versus XGBoost (AUROC 0.805, AUPRC 0.507), Logistic Regression (AUROC 0.795, AUPRC 0.524), and Random Forest (AUROC 0.801, AUPRC 0.520). Notably, AUPRC gains were most pronounced at the 90-day horizon, where class imbalance makes precision-recall performance a particularly stringent test of model utility.

**Figure 1:**
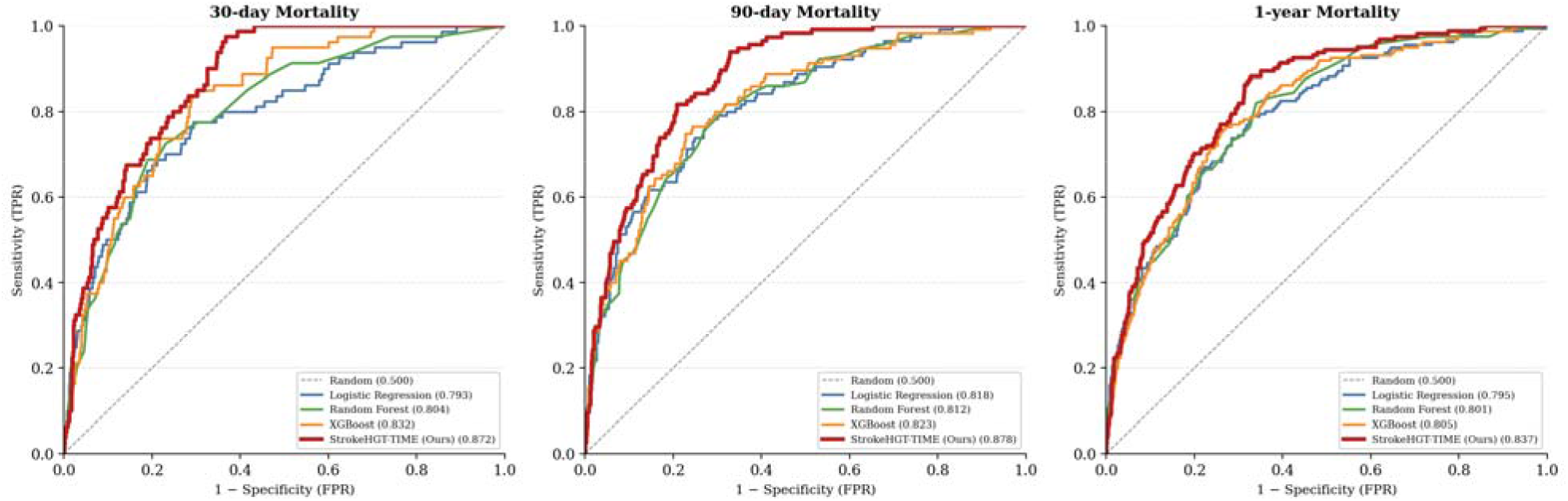
ROC curves for mortality prediction at 30 days, 90 days, and 1 year for proposed approach vs Baselines

**Figure 2:**
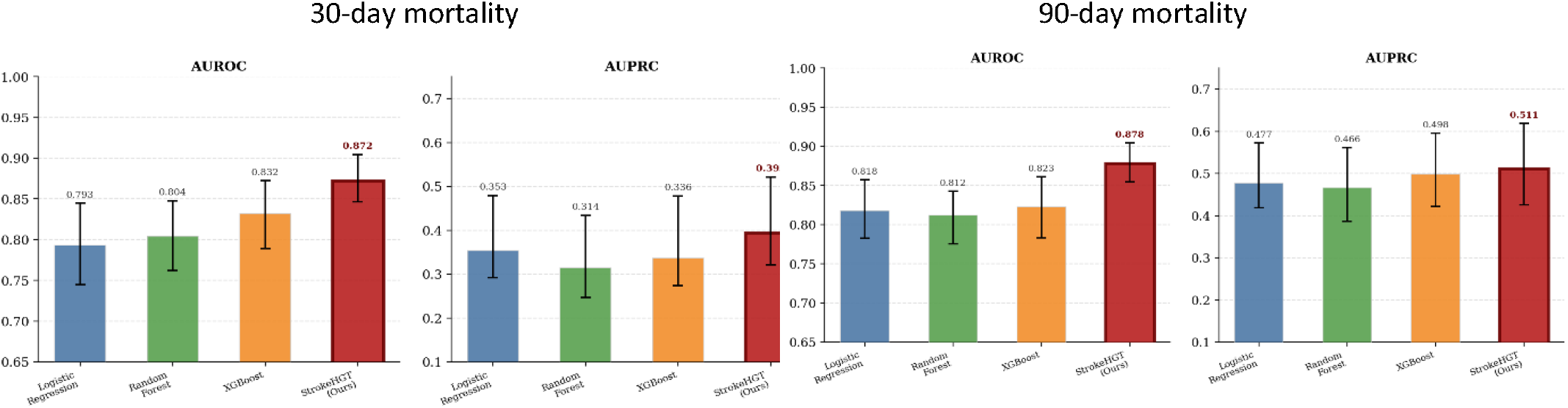
Performance comparison of proposed approach vs baseline

**Figure 3:**
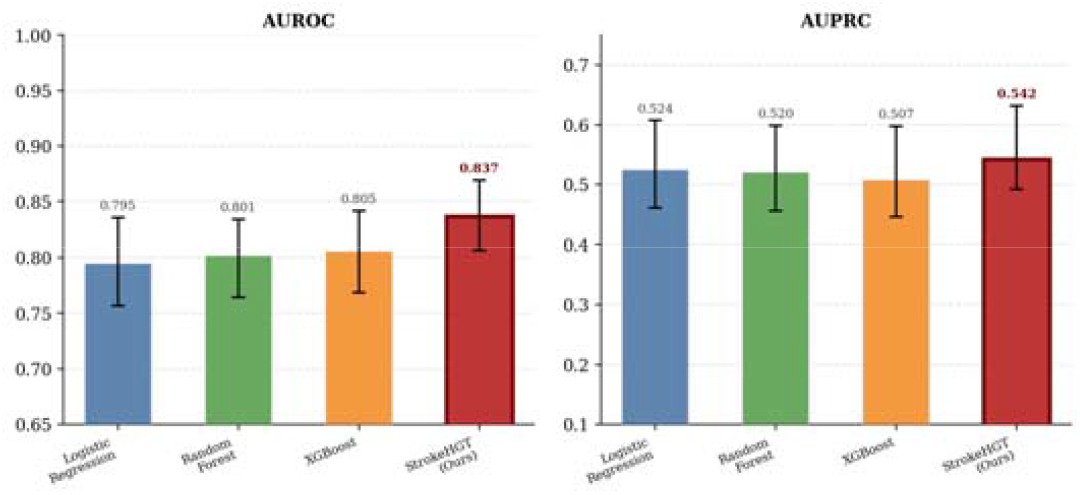
Performance comparison of proposed approach vs baseline for 1-year mortality

### Sensitivity at Fixed Specificity

Table 2 presents sensitivity and specificity for all models at a fixed specificity threshold of ≥75% across all three mortality horizons. StrokeTHG consistently identified more true mortality cases than all baselines while maintaining comparable specificity. At 30 days, StrokeTHG achieved a sensitivity of 0.787 at a realized specificity of 0.752, compared to XGBoost and Random Forest (both 0.738 sensitivity, specificity 0.732 and 0.748, respectively) and Logistic Regression (0.700 sensitivity, specificity 0.736). The advantage was most pronounced at 90 days, where StrokeTHG reached a sensitivity of 0.835, approximately 7.8 percentage points above XGBoost (0.757) and over 10 points above Logistic Regression (0.730) and Random Forest (0.722), while maintaining a specificity of 0.751, on par with all baselines. At one year, StrokeTHG achieved a sensitivity of 0.739 (specificity 0.750), outperforming XGBoost (0.720), Random Forest (0.689), and Logistic Regression (0.677).

**Table 2:** Sensitivity and specificity of all models across prediction horizons. All thresholds fixed at ≥75% specificity. Bold values indicate the best performance.

| Model | 30 day |  | 90 day |  | 1 year |  |
| --- | --- | --- | --- | --- | --- | --- |
|  | Sensitivity | Specificity | Sensitivity | Specificity | Sensitivity | Specificity |
| Logistic Regression | 0.700 | 0.736 | 0.730 | 0.752 | 0.677 | 0.760 |
| Random Forest | 0.738 | 0.748 | 0.722 | 0.744 | 0.689 | 0.741 |
| XGBoost | 0.738 | 0.732 | 0.757 | 0.756 | 0.720 | 0.747 |
| StrokeTHG | **0.787** | **0.752** | **0.835** | **0.751** | **0.739** | **0.750** |

### Representation Quality Analysis

To assess whether the performance gains of StrokeTHG originate from the quality of its learned representations rather than classifier choice, we conducted a controlled embedding comparison (Table 3). We evaluated four embedding methods: Non-negative Matrix Factorization (NMF)^*28*^, which captures linear co-occurrence structure in the patient-feature matrix; Spectral Embedding^*29*^ (SC), which encodes global graph topology via Laplacian eigenvectors; node2vec^*30*^, which learns node representations through random walks on a homogeneous patient similarity graph metapath2vec^*31*^, which extends random walks to heterogeneous graphs via predefined meta-paths and a multitask MLP, whose penultimate layer embedding serves as a neural tabular baseline. All embeddings were evaluated using an identical Logistic Regression downstream classifier, thereby isolating representation quality from modeling architecture.

**Table 3:** Embedding Comparison, AUROC by Method and Prediction Horizon. All embeddings evaluated using identical Logistic Regression classifier. Bold value indicate the best performance.

| Method | 30 d AUROC | 90 d AUROC | 1y AUROC |
| --- | --- | --- | --- |
| NMF | 0.816 | 0.827 | 0.823 |
| Spectral (SC) | 0.697 | 0.677 | 0.674 |
| node2vec | 0.762 | 0.792 | 0.787 |
| metapath2vec | 0.539 | 0.513 | 0.517 |
| MLP | 0.805 | 0.822 | 0.790 |
| StrokeTHG (Ours) | **0.84** | **0.852** | **0.821** |

Results revealed a clear but non-monotonic hierarchy of representation quality. NMF achieved competitive AUROCs of 0.816, 0.827, and 0.823 at 30-day, 90-day, and 1-year horizons, respectively. Spectral embedding underperformed across all horizons (AUROC: 0.697, 0.677, 0.674). node2vec improved upon spectral methods (AUROC: 0.762, 0.792, 0.787). metapath2vec collapsed to near-chance performance despite operating on the full heterogeneous graph (AUROC: 0.539, 0.513, 0.517). The multitask MLP achieved AUROCs of 0.805, 0.822, and 0.790 at 30-day, 90-day, and 1-year horizons respectively. StrokeTHG embeddings achieved the highest AUROC at the 30-day and 90-day horizons (0.840 and 0.852, respectively) and performed comparably to NMF at the 1-year horizon (0.821 vs. 0.823).

### Subgroup Performance Analysis

To evaluate the generalizability of StrokeTHG across clinically relevant patient strata, we stratified the held-out test set by age group and sex and assessed AUROC performance for all models within each subgroup.

#### Age Subgroups

Across all three age groups (Table 4), StrokeTHG demonstrated consistent discriminative advantages over baseline models, with performance gains varying systematically with age. In the Age <65 subgroup (n=303), StrokeTHG achieved the strongest absolute performance across all horizons, reaching an AUROC of 0.920 at 90 days, the highest value observed across any subgroup or model in this study, representing a +0.031 improvement over the best baseline (XGBoost: 0.887). At 30 days, the model achieved an AUROC of 0.890 (+0.036 over XGBoost: 0.854). In the Age 65–80 subgroup (n=343), StrokeTHG exhibited its largest margin improvements, with gains of +0.046 at 30 days (0.863 vs. Logistic Regression: 0.816) and +0.060 at 90 days (0.845 vs. Logistic Regression: 0.785).

**Table 4:** Subgroup Performance Analysis by Age Group, StrokeTHG vs Baselines (AUROC). Bold value indicate the best performance.

| Subgroup | Model | 30d AUROC | 90d AUROC | 1y AUROC |
| --- | --- | --- | --- | --- |
| Age <65 (n=303) | Logistic Regression | 0.84 | 0.889 | 0.821 |
|  | Random Forest | 0.853 | 0.872 | 0.85 |
|  | XGBoost | 0.854 | 0.887 | 0.836 |
|  | **StrokeTHG** | **0.890 (+0.036)** | **0.920 (+0.031)** | **0.861 (+0.010)** |
| Age 65–80 (n=343) | Logistic Regression | 0.816 | 0.785 | 0.776 |
|  | Random Forest | 0.778 | 0.767 | 0.762 |
|  | XGBoost | 0.809 | 0.77 | 0.726 |
|  | **StrokeTHG** | **0.863 (+0.046)** | **0.845 (+0.060)** | **0.804 (+0.028)** |
| Age >80 (n=183) | Logistic Regression | 0.8 | 0.845 | **0.787** |
|  | Random Forest | 0.779 | 0.769 | 0.715 |
|  | XGBoost | 0.82 | 0.802 | 0.71 |
|  | **StrokeTHG** | **0.830 (+0.010)** | **0.846 (+0.001)** | 0.776 (−0.011) |

Performance advantages were attenuated in the Age >80 subgroup (n=183), with near-parity at 90 days (+0.001) and a marginal deficit at the 1-year horizon, where Logistic Regression (0.787) marginally outperformed StrokeTHG (0.776, Δ = −0.011).

#### Sex Subgroups

StrokeTHG demonstrated comparable performance advantages across female (n=360) and male (n=469) subgroups (Table 5), suggesting no substantial sex-based differential in model performance, though larger and more demographically diverse cohorts would be needed to draw firm conclusions about representational fairness. Among female patients, AUROC improvements ranged from +0.015 at 1 year to +0.029 at 90 days, while in male patients, gains ranged from +0.005 at 1 year to +0.031 at 90 days. The 90-day horizon showed the largest sex-stratified gains for both groups, consistent with the full cohort results.

**Table 5:** Subgroup Performance Analysis by Sex, StrokeTHG vs Baselines (AUROC)

| Subgroup | Model | 30d AUROC | 90d AUROC | 1y AUROC |
| --- | --- | --- | --- | --- |
| Female (n=360) | Logistic Regression | 0.859 | 0.851 | 0.837 |
|  | Random Forest | 0.799 | 0.792 | 0.798 |
|  | XGBoost | 0.836 | 0.821 | 0.807 |
|  | **StrokeTHG** | **0.885 (+0.027)** | **0.879 (+0.029)** | **0.852 (+0.015)** |
| Male (n=469) | Logistic Regression | 0.818 | 0.84 | 0.814 |
|  | Random Forest | 0.831 | 0.845 | 0.818 |
|  | XGBoost | 0.846 | 0.847 | 0.781 |
|  | **StrokeTHG** | **0.864 (+0.019)** | **0.878 (+0.031)** | **0.823 (+0.005)** |

### Edge-Type Importance Analysis

To interpret which relational connections in the heterogeneous graph drive StrokeTHG’s mortality predictions, we conducted an edge-type ablation analysis. Each edge type was systematically removed from the graph, and the mean absolute change in predicted mortalit probability across all patients and horizons was recorded as an importance score (Figure 4). The most influential edge type was phenotype→patient (importance = 0.0288), followed by admission→patient (0.0249). Patient similarity edges ranked third and fourth patient→patient (similar_to, 0.0176) and patient→patient (lab_similar_to, 0.0151).

**Figure 4:**
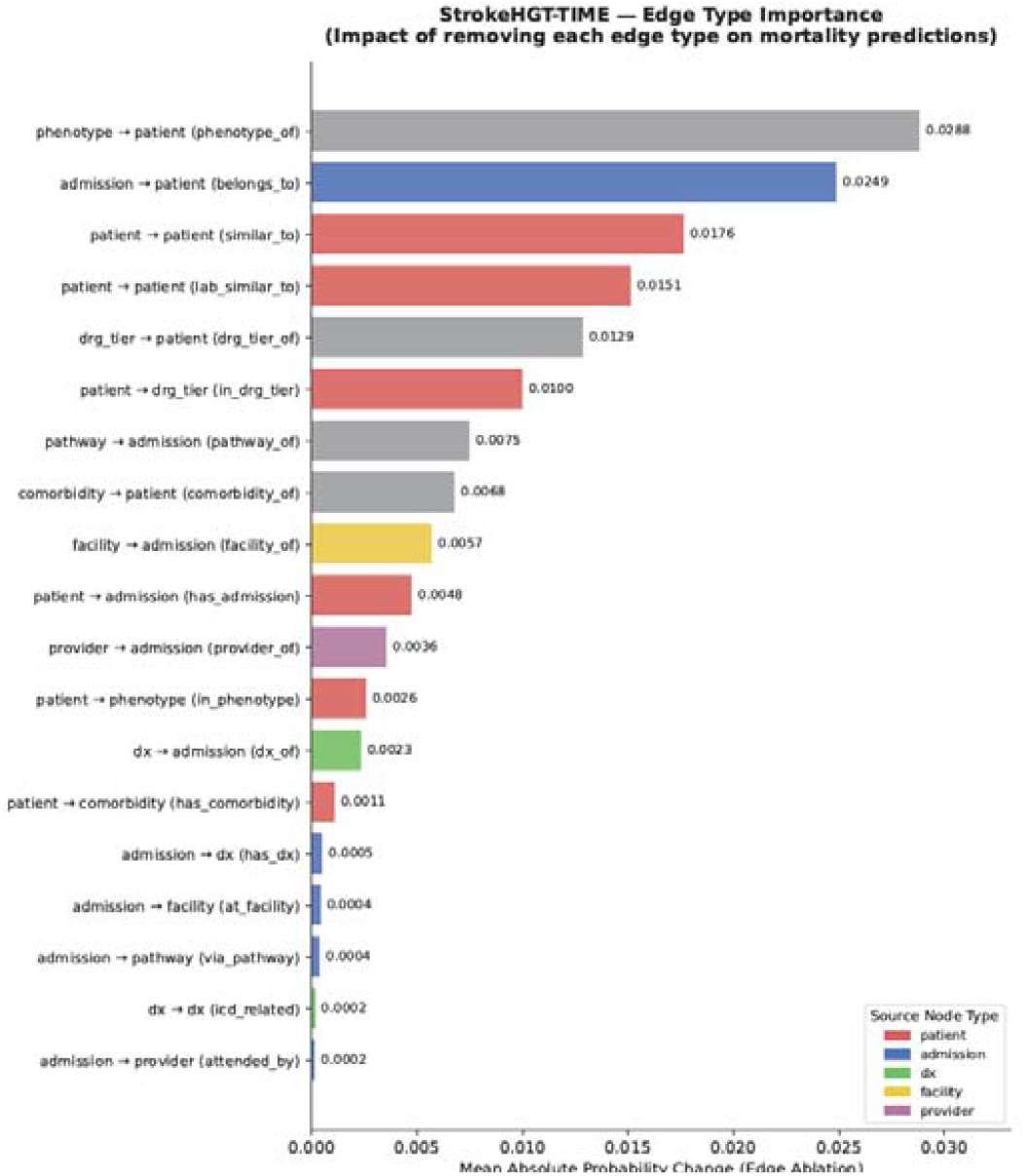
Edge type importance. Impact of removing each type on mortality predictions

## Discussion

This study presents StrokeTHG, a heterogeneous graph representation of EHR data. We demonstrate that heterogeneous graph representations of EHR data consistently improve multi-horizon stroke mortality prediction over flat tabular models across all three prediction horizons, supporting our central hypothesis that relational EHR structure encodes clinically meaningful signals beyond what tabular feature vectors can capture.

### Interpretation of Key Findings

At a fixed specificity of ≥75%, StrokeTHG identified 78.7%, 83.5%, and 73.9% of 30-day, 90-day, and 1-year mortality cases, respectively, consistently 5–10 percentage points above the best tabular baseline at each horizon. In a clinical deployment context, this translates directly to fewer missed high-risk patients per unit of clinical workload, which is arguably more important than raw AUROC for triage and early intervention workflows. These consistent improvements across both AUROC and AUPRC confirm that the heterogeneous graph structure provides incremental discriminative value beyond tabular feature representations.

The edge-type ablation analysis provides mechanistic insight into these gains. The phenotype→patient edge was the most influential connection, indicating that population-level phenotypic cluster memberships, derived from K-means clustering on laboratory profiles, carry the strongest relational signal for mortality prediction. This finding highlights the value of implicit collaborative filtering: the model effectively learns from the mortality outcomes of phenotypically similar patients, augmenting individual-level feature information with population-level risk patterns. The second most influential edge, admission→patient, underscores the importance of temporal admission history and its role in propagating longitudinal risk signals through the graph. The meaningful contribution of both patient-to-patient similarity edges, patient→patient (similar_to) and patient→patient (lab_similar_to), supports the design choice of incorporating both demographic/comorbidity-based and laboratory-based KNN channels, confirming that patient-to-patient relational structure provides a discriminative signal beyond individual node features. Together, these edge types account for the majority of the model’s relational reasoning and support the key architectural design choices in StrokeTHG, demonstrating that heterogeneous graph construction can surface latent clinical relationships that are not directly observable in flat tabular EHR representations.

The representation quality analysis further strengthens these findings. When evaluated under an identical Logistic Regression downstream classifier, StrokeTHG embeddings achieved the highest AUROC at the 30-day and 90-day horizons (0.840 and 0.852, respectively) and performed comparably to NMF at the 1-year horizon (0.821 vs. 0.823). NMF’s competitive performance reflects its ability to capture latent co-occurrence patterns across clinical features through low-rank matrix factorization. Spectral embedding’s underperformance suggests that graph geometry alone, without relational semantics, provides insufficient signal for mortality stratification. node2vec’s improvement over spectral methods is limited by its homogeneous formulation, which cannot exploit the multi-type relational structure inherent to clinical data. The collapse of metapath2vec to near-chance performance, despite operating on the full heterogeneous graph, likely reflects insufficient meta-path coverage across the sparse edge structure of the clinical graph, aligning with recent observations that pretraining on graph topology without outcome supervision yields limited downstream utility in sparse, noisy EHR graphs. The multitask MLP, which represents the upper bound of neural feature extraction without graph structure, was consistently outperformed by StrokeTHG embeddings across all three prediction horizons (ΔAUROC = +0.035, +0.030, and +0.031). Together, these findings confirm that StrokeTHG’s performance advantage is driven by the quality of its learned heterogeneous graph representations rather than downstream classifier capacity.

The subgroup analysis revealed an important pattern: performance advantages were largest in the Age 65–80 stratum (+0.046 at 30 days, +0.060 at 90 days) and smallest, and in one case reversed, in the Age >80 group (−0.011 at 1 year). This gradient likely reflects the differential informativeness of relational EHR structure across age strata. In middle-aged patients, a heterogeneous relational context, including comorbidity burden, care pathways, and phenotypic cluster membership, provides substantial discriminative lift over tabular features alone. The attenuation in the Age >80 subgroup likely reflects the higher baseline mortality prevalence and a more homogeneous risk profile in the oldest patients, where linear feature combinations may be sufficient for risk stratification, and the relational graph structure provides diminishing marginal benefit.

Across sex subgroups, performance gains were symmetric and consistent, with no substantial differential accuracy that would suggest sex-based representational bias. The symmetry of gains across sex subgroups, despite moderate sex imbalance in the test set (Women: n=360, Men: n=469), supports the equitable generalizability of the learned representations and the fairness of StrokeTHG for clinical deployment.

### Limitations and Future Work

Several limitations should be acknowledged. First, this study was conducted on a single institutional cohort from Penn State Health, and external validation on geographically and demographically diverse stroke populations is necessary before clinical deployment. The model’s performance in populations with different EHR coding practices, care pathways, or stroke subtype distributions remains unknown. Second, while the edge-type ablation analysis provides model-level interpretability, patient-level explanations, such as which specific graph neighborhoods or node connections drove an individual patient’s risk prediction, remain an important direction for future work to support clinical trust and adoption. Third, the heterogeneous graph was constructed using a general-purpose relational schema rather than a stroke-specific one. Node types and edge connections were defined based on broad EHR entity relationships, and a more systematic graph design process that encodes stroke-specific clinical signals, such as stroke severity markers, neuroimaging findings, reperfusion therapy status, and post-stroke complications, could improve the clinical grounding of the learned representations. Fourth, temporal information in the current graph is limited to admission time encoded via sinusoidal embeddings on admission nodes. Richer temporal modeling, through the addition of event nodes that capture serial laboratory measurements, medication changes, readmission events, and clinical deterioration episodes as time-stamped graph updates, would allow the model to capture longitudinal disease trajectories more effectively, particularly for the 1-year mortality prediction horizon, where post-discharge dynamics are most relevant. Finally, while KNN similarity edges are constructed in a leakage-safe manner, shared structural nodes such as phenotype clusters and comorbidity nodes create indirect 2-hop connections between validation and test patients through which input features can propagate. This is consistent with standard transductive graph learning practice, but means the evaluation is not fully inductive. A strictly inductive evaluation, in which test patients are completely isolated from the graph during training, would provide a stronger test of generalizability and is identified as a direction for future work. In future studies, we will also assess training stability across multiple random initializations to characterize model variance.

Future work should prioritize several directions. External multi-site validation is the most critical next step, including evaluation on the Geisinger cohort already available within this research program. Adopting an inductive graph learning framework such as GraphSAGE would address the transductive limitations of the current approach, enabling fully isolated test evaluation and fairer comparison with tabular baselines. A more systematic, stroke-specific graph-construction process, guided by clinical domain knowledge and stroke-outcome literature, could further improve the model’s relational signal quality. Expanding temporal graph representations through event-level node modeling would better capture the dynamic nature of post-stroke recovery and deterioration. Finally, integrating natural language processing of clinical notes would enrich the graph with unstructured clinical information, including signs, symptoms, and neurological assessments, that is currently absent from the structured EHR graph.

## Conclusion

We presented StrokeTHG, a temporal heterogeneous graph neural network that jointly predicts 30-day, 90-day, and 1-year post-stroke mortality from EHR data while enforcing monotonic probability ordering across horizons. Evaluated on Penn State Health stroke patients, StrokeTHG consistently outperformed tabular and graph-based baselines across all prediction horizons, with the most pronounced advantages at clinically actionable sensitivity thresholds, in the Age 65–80 stratum, and with equitable performance across sex subgroups. Edge-type ablation revealed that phenotypic similarity structures and admission history linkages are the primary drivers of the model’s relational reasoning, demonstrating that heterogeneous graph construction can surface latent clinical relationships not directly observable in flat tabular EHR representations. Representation quality analysis confirmed that these gains are attributable to task-aware heterogeneous graph representations rather than downstream classifier capacity. Taken together, these findings support the broader applicability of heterogeneous graph neural networks for multi-horizon clinical risk stratification, and establish a methodological foundation for graph-based mortality prediction in stroke care pending external validation.

## Data Availability

The data that support the findings of this study are not publicly available due to privacy restrictions. However, de-identified data may be made available upon reasonable request to the corresponding author, subject to approval from the Penn State Institutional Review Board (IRB) and the execution of a formal Data Use Agreement.

## Code Availability

Code for this work will be shared in a GitHub repository upon acceptance of this manuscript.

## Notes

**Funding:** This work was partially supported by the National Institutes of Health under Award Number R01NS128986.

### Competing Interest Statement

The authors have declared no competing interest.

### Author Declarations

IRB of Penn State gave ethical approval for this work

