## Supplemental material for "A Heterogeneous Graph Neural Network Framework for Multi-Horizon Stroke Mortality Prediction"

**Supplementary Material**

**Table S1** Patient Demographic and Clinical Characteristics (N=4,144)

| **Characteristic** | **Overall N=4,144** | **Survived (1y) N=3,332 (80.4%)** | **Died (1y) N=812 (19.6%)** |
| --- | --- | --- | --- |
| **Demographics** | | | |
| Age at index stroke, years — mean (SD) | 69.0 (14.7) | 67.1 (14.6) | 77.1 (12.2) |
| Male sex — n (%) | 2,252 (54.3%) | 1,861 (55.9%) | 391 (48.2%) |
| **Race/Ethnicity — n (%)** | | | |
| White | 3,438 (83.0%) | 2,729 (81.9%) | 709 (87.3%) |
| Black/African American | 278 (6.7%) | 235 (7.1%) | 43 (5.3%) |
| Multiracial | 195 (4.7%) | 161 (4.8%) | 34 (4.2%) |
| Asian | 72 (1.7%) | 68 (2.0%) | 4 (0.5%) |
| **Comorbidities — n (%)** | | | |
| Hypertension | 3,735 (90.1%) | 2,970 (89.1%) | 765 (94.2%) |
| Dyslipidemia | 3,496 (84.4%) | 2,853 (85.6%) | 643 (79.2%) |
| Diabetes mellitus | 1,780 (43.0%) | 1,399 (42.0%) | 381 (46.9%) |
| Atrial fibrillation | 1,582 (38.2%) | 1,124 (33.7%) | 458 (56.4%) |
| Heart failure | 1,104 (26.6%) | 781 (23.4%) | 323 (39.8%) |
| Chronic kidney disease | 1,011 (24.4%) | 747 (22.4%) | 264 (32.5%) |
| Chronic lung disease | 1,025 (24.7%) | 824 (24.7%) | 201 (24.8%) |
| Peripheral vascular disease | 901 (21.7%) | 711 (21.3%) | 190 (23.4%) |
| Myocardial infarction | 781 (18.8%) | 576 (17.3%) | 205 (25.2%) |
| Neoplasm | 736 (17.8%) | 532 (16.0%) | 204 (25.1%) |
| Atrial flutter | 258 (6.2%) | 189 (5.7%) | 69 (8.5%) |
| Hypercoagulable states | 235 (5.7%) | 173 (5.2%) | 62 (7.6%) |
| Rheumatic diseases | 215 (5.2%) | 177 (5.3%) | 38 (4.7%) |
| Mean comorbidities per patient | 4.1 | 3.9 | 4.7 |

**Table S2** ICD codes for ischemic stroke identification.

| **Code system** | **No. of codes** | **Code prefixes / examples** |
| --- | --- | --- |
| ICD-9 | 9 | 433.x1 (e.g. 433.01, 433.11), 434.x1 (e.g. 434.01, 434.11) |
| ICD-10 | 118 | H34.1; I63 family (I63.0–I63.9, including sub‐codes such as I63.011, I63.312, etc.) |

**Table S3** Model Hyperparameters and Training Details - StrokeTHG

| **Hyperparameter** | **Value** |
| --- | --- |
| Hidden dimension | 128 |
| Message passing layers | 3 |
| Attention heads | 4 |
| Dropout | 0.35 |
| Stochastic depth (drop_path) | 0.05 |
| Head hidden dimension | 64 |
| Optimizer | AdamW |
| Learning rate | $3\times10⁻⁴$ |
| Weight decay | $10⁻⁴$ |
| Max epochs | 500 |
| Early stopping patience | 60 evaluation steps |
| Evaluation frequency | Every 5 epochs |
| Early stopping metric | Mean validation AUROC across 3 horizons |
| Gradient clipping | Norm 2.0 |
| Loss function | Asymmetric focal loss (γ₊=1.0, γ₋=3.0, label smoothing=0.05) |
| Pre-training epochs | 150 (masked feature reconstruction, 30% masking rate) |
| Random seed | 42 |

**Table S4** Model Hyperparameters and Training Details - Baseline Models

| **Model** | **Hyperparameter** | **Value** |
| --- | --- | --- |
| Logistic Regression | Regularization | L2 |
|  | C | 1 |
|  | Max iterations | 1000 |
|  | Solver | saga |
|  | Class weight | balanced |
| Random Forest | n_estimators | 100 |
|  | Max depth | default (None) |
|  | Class weight | balanced |
|  | n_jobs | -1 |
| XGBoost | n_estimators | 300 |
|  | Learning rate | 0.05 |
|  | Max depth | 5 |
|  | Subsample | 0.8 |
|  | Scale pos weight | n_neg/n_pos per horizon |
|  | eval_metric | logloss |

**Table S5** Training details (all models)

| **Detail** | **Value** |
| --- | --- |
| Train/Val/Test split | 60/20/20 stratified by 30-day mortality |
| Feature standardization | StandardScaler fit on training set only |
| Confidence intervals | BCa bootstrap (500 iterations) |
| Framework | PyTorch 2.8.0, PyTorch Geometric 2.5.2, scikit-learn |

**Table S6** Paired Bootstrap AUROC Difference Analysis, StrokeTHG vs Baseline Models

Penn State Health Test Set (N=829), 10,000 Bootstrap Resamples

| **Horizon** | **Comparison** | **HGT AUROC** | **Baseline AUROC** | **ΔAUROC** | **95% CI Lower** | **95% CI Upper** | **p-value** | **Significant** |
| --- | --- | --- | --- | --- | --- | --- | --- | --- |
| **30-day** | StrokeTHG vs Logistic Regression | 0.872 | 0.793 | 0.035 | 0.008 | 0.062 | 0.008 | Yes* |
|  | StrokeTHG vs Random Forest | 0.872 | 0.804 | 0.058 | 0.02 | 0.098 | 0.002 | Yes* |
|  | StrokeTHG vs XGBoost | 0.872 | 0.832 | 0.03 | −0.004 | 0.064 | 0.081 | No |
| **90-day** | StrokeTHG vs Logistic Regression | 0.878 | 0.818 | 0.034 | 0.01 | 0.059 | 0.004 | Yes* |
|  | StrokeTHG vs Random Forest | 0.878 | 0.812 | 0.058 | 0.027 | 0.09 | <0.001 | Yes* |
|  | StrokeTHG vs XGBoost | 0.878 | 0.823 | 0.045 | 0.019 | 0.072 | 0.001 | Yes* |
| **1-year** | StrokeTHG vs Logistic Regression | 0.837 | 0.807 | 0.013 | −0.012 | 0.037 | 0.32 | No |
|  | StrokeTHG vs Random Forest | 0.837 | 0.808 | 0.027 | −0.001 | 0.056 | 0.06 | No† |
|  | StrokeTHG vs XGBoost | 0.837 | 0.816 | 0.042 | 0.013 | 0.07 | 0.004 | Yes* |
